# Are COVID-19 infected children with gastrointestinal symptoms different from those without symptoms? A comparative study of the clinical characteristics and epidemiological trend of 244 pediatric cases from Wuhan

**DOI:** 10.1101/2020.04.29.20084244

**Authors:** Xiao-li Xiong, Kenneth Kak-yuen Wong, Shui-qing Chi, Ai-fen Zhou, Jian-qiao Tang, Li-Shan Zhou, Patrick Ho-yu Chung, Gilbert T Chua, Keith TS Tung, Ian CK Wong, Celine SL Chui, X Li, Mike Yat-wah Kwan, Wilfred Hing-sang Wong, Marco Hok-kung Ho, Godfrey Chi-fung Chan, Guoqing Cao, Kang Li, Patrick Ip, Peng Chen, Shao-tao Tang, Paul Kwong-hang Tam

## Abstract

**Objective:** COVID-19 patients presenting with gastrointestinal (GI) symptoms occur in both adults and children. To date, however, no large sample size study focusing on gastrointestinal symptoms in pediatric cases has been published. We analyzed COVID-19 infected children in Wuhan who presented with initial GI symptoms to determine the GI characteristics and epidemiological trend of the disease.

**Design:** We retrospectively analyzed 244 children patients confirmed with COVID-19 at Wuhan Children’s Hospital from 21 Jan to 20 Mar 2020. Symptomatic cases were divided into two groups according to whether the patients presented with or without GI symptoms on admission. Demographic, epidemiological, symptoms, and laboratory data were compared. We also analyzed the respective trends of case number changes of GI cases and asymptomatic cases.

**Results:** 34 out of 193 symptomatic children had GI symptoms. They had lower median age and weight, a higher rate of fever, a longer length of stay and more hematological and biochemical abnormalities than patients without GI symptoms. There was no significant difference in chest CT findings or stool SARS-CoV-2 test positive percentages between the two groups. The number of patients admitted with GI symptoms showed an overall downward trend with time. At the time of writing, 242 patients were discharged, one died, and one critically ill patient was still in the intensive care unit.

**Conclusion:** COVID-19 infected children with GI symptoms are prone to presenting with more clinical and laboratory abnormalities than patients without GI symptoms. More attention and timely hospital admission are needed for these patients.

**Significance of this study:**

What is already known on this subject?
- COVID-19 is now a pandemic with more than 1.6 million people infected worldwide
- Although attacking the respiratory tract mostly, both adult and children infected with COVID-19 can present with GI symptoms

What are the new findings?
- Infants younger than two years old and presence of fever are the two risk factors of presenting with GI symptoms
- A high proportion of patients without GI symptoms and asymptomatic patients will have positive RT-PCR for the virus in stool
- Earlier testing through contact screening of family members means more COVID-19 infected children are diagnosed when completely asymptomatic

How might it impact on clinical practice in the foreseeable future?
- The presence of COVID-19 in stool in infected children will have a major implication for parents and carers of young infants
- Increasing number of asymptomatic COVID-19 patients who are detected through screening could provide a useful lesson for other countries still experiencing the rise and peak of the pandemic

## Introduction

COVID-19 was first confirmed in Wuhan, Hubei province in December 2019, and has rapidly become a global disease of epic scale in less than two months. As of to date, this pandemic has still not been well controlled, [1, 2, 3] Similar to the Severe Acute Respiratory Syndrome (SARS) in 2003, apart from respiratory manifestations, a proportion of COVID-19 patients also develop GI symptoms, such as diarrhea and vomiting. [4, 5] Several retrospective studies have described the clinical symptoms, epidemiologic characteristics, and outcomes of the patients with GI symptoms, but these have mainly focused on the adult population. [6, 7, 8, 9] It is still not known whether children with COVID-19 have similar or different GI features as adults. Furthermore, some recent studies reported the identification of asymptomatic COVID-19 patients. This has raised the possibility and risk of disease transmission by these patients as silent carriers. [10, 11] In this cohort, some patients were diagnosed as asymptomatic infection as well. The main purpose of this study was to describe the gastrointestinal clinical characteristics, to compare the differences between patients with and without GI symptoms, and to find out the epidemiological trend of COVID-19 children in Wuhan.

## Methods

### Study design and patients

This was a retrospective single-center study approved by the Research Ethics Board of Wuhan Children’s Hospital (**WHCH2020022**). Between 21 Jan 2020 and 20 Mar 2020, 244 patients with positive real-time reverse transcription-polymerase chain reaction (RT-PCR) results for COVID-19 from the laboratory department or Wuhan Center for Disease Control and Prevention for nasopharyngeal swab specimens were included. For patients confirmed to have COVID-19, they were divided into five sub-types, according to The Second Edition of Children’s COVID-19 Infection Diagnosis, Treatment, and Prevention Guidelines:[12]

1. Asymptomatic - patients without any symptom or abnormal radiography finding.
2. Acute upper respiratory tract infection - patients presenting with fever, cough, sore throat, fatigue, or other symptoms without pneumonia symptoms on radiography examinations.
3. Mild pneumonia type - patients diagnosed when computed tomography (CT) of the thorax showed any finding of viral pneumonia.
4. Severe type - patients meeting at least one of the following criteria: i) Precluding the influence of high fever and crying, respiratory rate (RR) ≥ 60/min (for children <2 months old); RR ≥ 50/min (2–12 months old); RR ≥ 40/min (1–5 years old); RR ≥ 30/min (>5 years old); ii) Resting oxygen saturation ≤ 92%; iii) Dyspnea; iv) Disturbance of consciousness, such as drowsiness or convulsions; v) Dehydration symptoms; vi) High-resolution CT showing rapidly progressive pneumonia or pleural effusion.
5. Critically ill patients - patients meeting any of the following criteria: i) respiratory failure which requires mechanical ventilation; ii) septic shock; iii) multiple organ dysfunction syndromes (MODS).

Symptomatic patients were divided into two groups according to whether the patients had GI symptoms at presentation or not. Patients with GI symptoms were those having at least one of the following before admission: diarrhea (passing of loose stools >3 times per day); nausea and vomiting; abdominal pain; and anorexia. The medical records of all the patients were reviewed. The presenting symptoms, clinical classifications, nucleic acid positive duration, laboratory data, radiography reports, admission date, clinical progress and length of stay (LOS) were recorded. Patients were only discharged when their nasopharyngeal swab RT-PCR results turned negative at least twice on consecutive days.

### Patient and Public Involvement

Patients or the public WERE NOT involved in the design, or conduct, or reporting, or dissemination plans of our research

### Statistical analysis

Categorical variables were described as frequencies and percentages; continuous variables were described as mean (standard deviation) or median (interquartile range) according to the distribution of the data. Significant differences between two groups were tested by Pearson’s Chi-squared test for categorical variables, and Mann–Whitney U test or Student t-test for continuous variables. Univariate analysis for odds to presenting GI symptoms was performed by logistic regression for possible confounders from the comparison of patients with or without GI symptoms. Then a multivariable logistic regression model was built considering all significant variables from the univariate regression. Results were presented as odds ratio (OR) [95% confidence interval]. The data was analyzed by SPSS 26.0, and P<0.05 was used to determine significance.

## Results

Of the 244 children, 51 patients were asymptomatic. Thus, 193 symptomatic patients were divided into two groups according to whether they had GI symptoms or not on admission. Thirty-four patients (17.7%) presented with GI symptoms, and the other 159 patients (82.3%) did not have any GI symptoms on admission (Figure 1). Many patients indeed presented with more than one GI symptoms: 23 (9.47%) had vomiting, 15 (6.17%) had diarrhea, 8 (3.29%) had anorexia, and 4 (1.65%) patients had abdominal pain.

**Figure 1.**
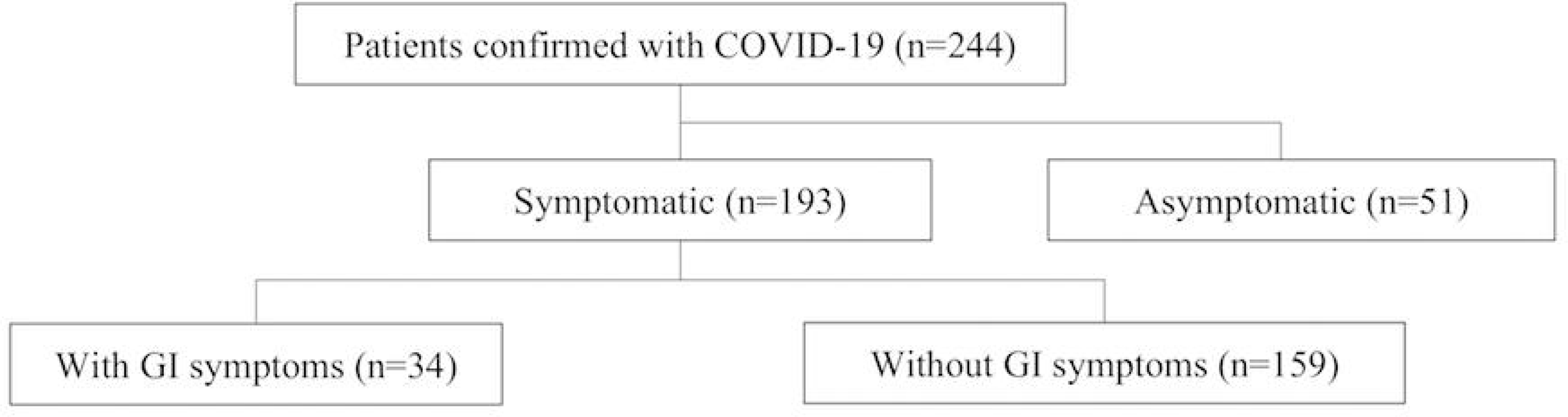
**–** A flowchart showing the cohort of 244 paediatric patients infected with COVID-19.

The two groups of patients were similar with respect to sex, birth weight, the percentage of patients in each COVID-19 clinical classification, and the percentage with contact history with an infected family member. The median of age at presentation and the age distribution were significantly different between the two groups. The patients with GI symptoms were much younger (14 vs. 74 months; P<0.05), and more than half were under three years old (Table 1).

**Table 1.** Demographics and epidemiological characteristics of COVID-19 children with or without GI symptoms.

|  | GI symptoms (n=34) | Non-GI symptoms (n=159) | P-value |
| --- | --- | --- | --- |
| **Age (months)** |  |  |  |
| Median (IQR) | 14(3-93) | 74(13-122) | 0.013* |
| **Age group distribution** |  |  | 0.020* |
| 0- 1 month | 2 (5.88%) | 8 (5.03%) |  |
| 1 month-12 months | 14 (41.18%) | 29 (18.24%) |  |
| 1-3 years old | 4 (11.76%) | 20 (12.58%) |  |
| 3-6 years old | 3 (8.82%) | 19 (11.95%) |  |
| 6-10 years old | 5 (14.71%) | 43 (27.04%) |  |
| 10-18 years old | 6 (17.65%) | 40 (25.16%) |  |
| **Sex (male)** | 19 (55.88%) | 101 (63.52%) | 0.404 |
| **Birth Weight (kg)** | 3.30 (2.93-3.70) | 3.30 (3.00-3.55) | 0.948 |
| **Clinical Diagnostic Classification** |  |  | 0.465 |
| Acute upper respiratory infection | 7 (20.59%) | 43 (27.04%) |  |
| Mild Pneumonia | 25 (73.53%) | 107 (67.30%) |  |
| Severe Pneumonia | 0 (0%) | 7 (4.40%) |  |
| Critical Pneumonia | 2 (5.88%) | 2 (1.26%) |  |
| **Contact History with Infected Family Member** | 27 (79.41%) | 132 (83.02%) | 0.616 |

We then compared the clinical and laboratory parameters between the two groups of patients. For the panel of non-GI symptoms, the only significant difference found was that patients with GI symptoms were more likely to have fever on admission (70.6% vs. 46.2%, P<0.05). Although the LOS was slightly longer in the GI group, (12.5 vs. 11.5 days), this did not reach statistical significance. No other significant differences were found between the two groups in other symptoms. Neither the duration of RT-PCR positivity for COVID-19, nor computer tomography (CT) of the thorax was significant between the two groups (Table 2.1). One result of interest was the high rate of positivity of stool SARS-CoV-2 RT-PCR, even in patients without any GI symptoms (37.1%) and asymptomatic patients (31.6%).

**Table 2.1.** Clinical and radiological characteristics of **COVID-19** children with or without GI symptoms

|  | GI symptoms (n=34) | Non-GI symptoms (n=159) | P-value |
| --- | --- | --- | --- |
| **Symptoms** |  |  |  |
| Fever | 24 (70.59%) | 75 (46.17%) | 0.013* |
| Cough | 17 (50.00%) | 103 (64.78%) | 0.107 |
| Sputum Production | 2 (5.88%) | 23 (14.47%) | 0.176 |
| Runny Nose | 4 (11.76%) | 20 (12.58%) | 0.896 |
| Fatigue | 1 (2.94%) | 7 (4.40%) | 0.698 |
| Dyspnea | 2 (5.88%) | 9 (5.66%) | 0.960 |
| Sore Throat | 2 (5.88%) | 8 (5.03%) | 0.839 |
| Headache | 4 (11.76%) | 6 (3.77%) | 0.056 |
| Dizziness | 1 (2.94%) | 2 (1.26%) | 0.471 |
| Myalgia | 3 (8.82%) | 6 (3.77%) | 0.205 |
| Chest Distress | 2 (5.88%) | 9 (5.66%) | 0.960 |
| Edema | 0 (0%) | 2 (1.26%) | 0.511 |
| Convulsion | 1 (2.94%) | 1 (0.63%) | 0.227 |
| Chills | 0 (0%) | 2 (1.26%) | 0.511 |
| **Length of Stay (days)** | 12.50 (8.00-17.00) | 11.5 (8.75-16.00) | 0.646 |
| **Nucleic acid Positive Duration (days)** | 7.00 (5.00-10.00) | 7.00 (5.00-11.00) | 0.731 |
| **Stool SARS-CoV-2 RT-PCR Results** |  |  |  |
| Positive | 7/17(41.18%) | 26/70(37.14%) | 0.759 |
| **Imaging** |  |  |  |
| **Pulmonary Lesions** |  |  | 0.287 |
| Null | 9 (26.47%) | 46 (28.93%) |  |
| Unilateral | 11 (32.35%) | 68 (42.77%) |  |
| Bilateral | 14 (41.18%) | 45 (28.30%) |  |
| **Ground-glass Opacities** | 6 (17.65%) | 24 (15.09%) | 0.762 |

For the blood tests done on admission, which included complete blood picture, liver and renal function tests and clotting profile, the only statistical differences seen were in neutrophil count, total protein, albumin, ALT, AST and creatinine. However, all these values were still within the normal reference range. The same was also true for cytokine profile and inflammatory markers (Table 2.2).

**Table 2.2.** Laboratory characteristics of **COVID-19** children with or without GI symptoms

|  | GI symptoms (n=34) | Non-GI symptoms (n=159) | P-value |
| --- | --- | --- | --- |
| **Complete blood picture** |  |  |  |
| White blood cell count (×10<sup>9</sup>/L) | 6.97 (5.39-9.20) | 6.75 (5.37-8.20) | 0.642 |
| Hemoglobin (g/L) | 122.00 (107.00-130.50) | 125.50 (117.75-133.00) | 0.051 |
| Platelets (×10<sup>9</sup>/L) | 278.00 (200.00-376.00) | 288.00 (241.00-368.00) | 0.502 |
| Neutrophils (×10<sup>9</sup>/L) | 1.92 (1.07-2.98) | 2.61 (1.84-3.77) | 0.023* |
| Monocytes (×10<sup>9</sup>/L) | 0.48 (0.35-0.74) | 0.42 (0.35-0.56) | 0.146 |
| Lymphocytes (×10<sup>9</sup>/L) | 3.49 (2.11-5.26) | 2.95 (1.97-4.04) | 0.168 |
| **Liver Function test** |  |  |  |
| Total Bilirubin (μmol/L) | 6.70 (5.10-8.55) | 7.70 (5.40-9.90) | 0.265 |
| Direct Bilirubin (μmol/L) | 1.80 (1.30-2.60) | 2.20 (1.60-3.30) | 0.055 |
| Total Protein (g/L) | 63.60 (57.90-72.60) | 68.80 (64.30-72.40) | 0.038* |
| Albumin (g/L) | 43.90 (40.70-46.55) | 45.10 (43.10-47.70) | 0.018* |
| Globulin (g/L) | 21.01±5.85 | 23.29±5.51 | 0.756 |
| Alanine aminotransferase (U/L) | 22.00 (13.75-31.75) | 15.00 (11.00-28.00) | 0.015* |
| γ-glutamyl transpeptidase (U) | 14.00 (8.00-31.00) | 11.00 (9.00-16.00) | 0.261 |
| Aspartate aminotransferase (U/L) | 42.50 (26.00-62.25) | 30.00 (23.00-42.00) | 0.002* |
| alkaline phosphatase (U) | 186.00 (148.00-265.00) | 194.00 (157.00-244.00) | 0.745 |
| **Renal Function test** |  |  |  |
| Blood urea nitrogen (mmol/L) | 3.85 (2.89-4.60) | 4.00 (3.20-4.68) | 0.656 |
| Creatinine (μmol/L) | 24.25 (19.10-38.08) | 33.20 (25.85-45.10) | 0.006* |
| **Coagulation Function** |  |  |  |
| Prothrombin Time (s) | 10.70 (10.40-11.35) | 10.90 (10.60-11.38) | 0.384 |
| PT-INR | 0.91 (0.88-0.96) | 0.93 (0.89-0.97) | 0.191 |
| D-Dimer (mg/L FEU) | 0.27 (0.14-0.63) | 0.22 (0.13-0.36) | 0.289 |
| Antithrombin (%) | 112.30 (103.90-121.80) | 116.90 (106.80-123.20) | 0.221 |
| Fibrinogen (g/L) | 2.15 (1.84-2.34) | 2.15 (1.87-2.75) | 0.267 |
| Activated Partial Thromboplastin Time (s) | 30.80 (28.40-38.30) | 30.85 (28.70-34.00) | 0.489 |
| Thrombin Time (s) | 18.00 (17.60-18.85) | 18.30 (17.43-19.38) | 0.614 |
| **Cytokines** |  |  |  |
| IL-2 (pg/ml) | 1.46 (1.26-1.82) | 1.43 (1.22-1.70) | 0.705 |
| IL-4 (pg/ml) | 2.66 (2.16-3.24) | 2.62 (2.10-3.27) | 0.876 |
| IL-6 (pg/ml) | 5.08 (2.95-14.31) | 3.92 (2.95-6.67) | 0.117 |
| IL-10 (pg/ml) | 5.21 (3.37-8.81) | 3.80 (3.23-4.58) | 0.018* |
| TNF-α(pg/ml) | 1.83 (1.59-2.22) | 1.62 (1.21-2.19) | 0.097 |
| IFN-γ (pg/ml) | 3.65 (2.86-8.57) | 2.89 (2.35-4.21) | 0.073 |
| **Infection-related Biomarkers** |  |  |  |
| Procalcitonin (ng/ml) | 0.07(0.04-0.11) | 0.05 (0.04-0.08) | 0.125 |
| C-reactive protein(mg/l) | 0.75(0.75-5.05) | 0.75 (0.75-2.97) | 0.597 |
| Erythrocyte sedimentation rate (mm/h) | 6.50 (3.50-14.50) | 10.00 (3.00-19.00) | 0.437 |

At the time writing, 242 patients were discharged. One patient died of viraemia and multi-organ failure, one initially critically ill patient was still in the intensive care unit.

In order to ascertain what factors on admission could predict GI symptoms in COVID-19 patients, we employed univariate analysis. This identified several indicators: age; fever, neutrophils, AST, and IL-10. However, when we probed further using multivariable analysis, the only factors which were significant were age and presence of fever (Table 3).

**Table 3.** Univariate and multivariate logistic regression for odds to GI symptoms

| **Logistic Regression for odds to GI symptoms** | **Univariate OR (CI 95%)** | **P-value** | **Multivariable OR (CI 95%)** | **P-value** |
| --- | --- | --- | --- | --- |
| Age (>24 months old vs. <24 months old) | 3.209 (1.501-6.860) | 0.003* | 3.208(1.163-8.852) | 0.024* |
| Fever (no vs. yes) | 2.688(1.207-5.987) | 0.016* | 3.638(1.219-10.858) | 0.021* |
| Neutrophils count (reference normal range) | 0.275 (0.107-0.705) | 0.007* | 0.878(0.239-3.223) | 0.844 |
| AST (reference normal range) | 3.584(1.548-8.302) | 0.003* | 2.338(0.661-8.276) | 0.187 |
| IL-10 (reference normal range) | 4.016(1.618-9.967) | 0.003* | 1.527 (0.505-4.623) | 0.453 |

To review the overall trend of our patients with GI symptoms, we plotted our whole cohort according to the admission dates. The median admission date was 18^th^ February. There were significantly more GI cases in patients admitted before this date than after (26 vs. 7; P<0.001). Conversely, the patients admitted after 18^th^ February were mostly asymptomatic cases (47 vs. 4; P<0.001) (Figure 2).

**Figure 2.**
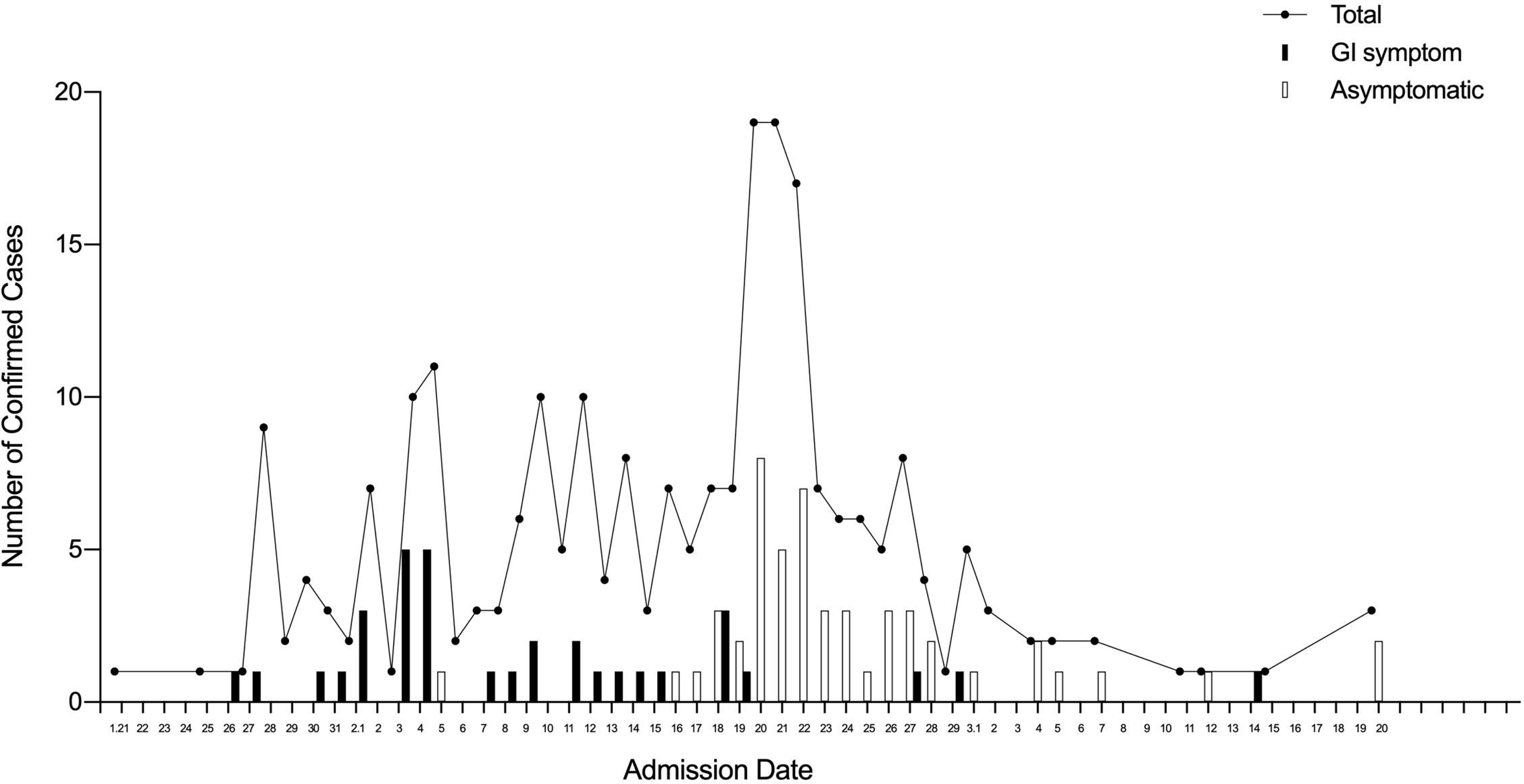
– A graph showing the total number of paediatric patients (line), the number of patients admitted with GI symptoms (solid bar) and the number of patients admitted without GI symptoms (blank bar) infected with COVID-19 between 21^st^ January to 20^th^ March 2020.

## Discussion

Several previous studies showed that GI symptoms were important in COVID-19 patients, and the fecal-oral route could be a possible route of transmission by SARS-CoV-2.[13] Furthermore, both adult and paediatric patients have been reported to have prolonged viral shedding in the faeces even when asymptomatic.[14,15] Thus, they will have an impact on an increased prevalence of this pandemic.[6, 16] To date, there is no large sample size study examining the impact of GI symptoms in children with COVID-19 and this study is to our knowledge the first to compare specifically paediatric patients with GI symptoms with those without GI symptom.

In our cohort, 17.7% presented with GI symptoms on admission, which is slightly higher than the 11.6% in an adult series.[7] While diarrhea was the most common complaint in adults (24.2%), vomiting (9.47%) was the predominant symptom, in children.[6, 7, 9] The reason for the higher rate of GI symptoms in children may be the difficulty in maintaining good hand hygiene in children who are more likely to touch their lips and mouth than adults. Supporting this, we found that infants (younger than two years old) were more likely to present with GI symptoms than older children. One more possible explanation for the higher incidence in younger children may be their less mature systemic and intestinal immunity.[17]

A study focusing on GI symptoms in COVID-19 adults noted that GI cases had a significantly higher rate of fever and a higher tendency to have a more severe disease.[6] In the current study for children, the results further support the association with fever, but not in terms of severity. The difference may be due to a small sample size in children and the extremely low incidence of severe COVID-19 cases in children, which was only 1.65% in this cohort. The presence of GI symptoms in children did not translate to significantly longer length of stay.

For laboratory abnormalities mainly concentrated on liver function and cytokines tests, significant abnormalities, including increases in AST, IL-10 and decreases in neutrophil, total protein and albumin were identified. Similar abnormalities had been described in the severe or critically ill case series reported previously.[18] These findings suggested that patients with GI symptoms might have more active inflammation especially in the liver. The absence of any significant difference in fecal nucleic acid RT-PCR between children with or without GI symptoms is in agreement with the findings of the adult cohort of Lin et al.[7] In fact, a high proportion of asymptomatic children were found to have positive RT-PCR for the virus in stool.

Although the detection of viral RNA without additional virological evidence, such as culture or detection of anti-genomic RNA, does not necessary imply infection, this nonetheless should alert us that contact precaution should be exercised in dealing with the excreta of patients with SARS-CoV-2, no matter that they have GI symptoms or not.

Several studies suggested that there might be an increasing trend in adults to present with GI symptoms, from about 3% reported in January to over 10% in February.[4, 8] However, our study in children showed a reverse trend. Three-quarters of the patients with GI symptoms were admitted before the median admission date.

This means over the course of the epidemic, fewer patients developed GI symptoms and more asymptomatic patients were diagnosed with passage of time. Whether it is related to the change of viral virulence or isolation policy remains to be further explored. Another observation is that most of the children were infected via family contact rather than community acquired.[16] In our cohort, 203 (83.1%) of 244 patients were infected by their parents or grandparents. As the epidemic progressed in Wuhan, diagnostic tests became more readily available leading to earlier disease detection through contact screening of family members before they became symptomatic. This observation could be useful for public health planning in countries still experiencing the early stage of the pandemic.

There are limitations of this study. This is a retrospective cohort study so we may not be able to establish any causal relationship. The sample size of patients with GI symptoms is relatively small, due to its low incidence so we may miss out some risk factors for its occurrence. The laboratory examinations and fecal SARS-CoV-2 RNA detection were not done in all patients and this may generate bias in the data analysis. Our study is one of the largest of its kind, and we showed the differences between clinical and laboratory characteristics of patients with or without GI symptoms, which were different from adults. Both pediatricians and patients need to pay more attention to COVID-19 children with GI symptoms even though they have mild symptoms and stable vital signs. An upward trend in asymptomatic infections also means the public should be on guard at all times.

## Data Availability

The availability of all data referred to in the manuscript can be obtained via the corresponding authors

## Competing Interests Declaration

All authors declare no competing interests.

## Notes

### Competing Interest Statement

The authors have declared no competing interest.

### Funding Statement

No external funding was received

